# Trauma under the influence: A review of Drug and Alcohol-related Major Trauma in North Queensland

**DOI:** 10.64898/2025.12.19.25342610

**Authors:** Michael Ryan, Breeanna Spring, Benjamin J Crowley, Bryannii Perks, Anna Grant, Christiaan Pretorius, Matan Ben David, Andrew Amos

## Abstract

**Objective:** Alcohol and drug use is an established factor contributing to traumatic injury globally. However, its prevalence and impact within regional and remote populations is poorly characterised. This study examines associations between alcohol and drug use and clinical outcomes among major trauma patients in North Queensland.

**Methods:** A retrospective audit was conducted of all major trauma patients in the Townsville University Hospital Trauma Registry Information System between October 2022 and October 2023. Demographic, injury and outcome data were investigated. Patients were categorised by alcohol and drug use and compared using descriptive and non-parametric analysis.

**Results:** Of 319 eligible patients, 110 (34.4%) patients were identified within the alcohol and drug cohort. Alcohol was most frequently detected (82.7%) followed by cannabis (20.0%) and amphetamines (12.7%) and 20 (18.2%) reported polysubstance use. Alcohol and/or drug-positive patients were significantly younger (40.7 vs 49.0 years, p<0.05), more likely to be injured by assault (OR 6.9 (95% CI: 3.1-15.3), p<0.001), admitted to ICU (OR 1.5 (95% CI: 0.9-2.5), p=0.069) and required longer hospital stays (11.9 ± 13.9 days vs 10.4 ± 11.4 days, p=0.656). Discharge and mortality outcomes did not differ between groups.

**Conclusion:** This study highlights the impact of alcohol and drug use in regional major trauma patients, who often require greater healthcare utilisation. Standardised testing, targeted prevention and tailored clinical pathways may improve outcomes and reduce the burden of alcohol and drug-related trauma. Due to the inconsistent screening practices, the true prevalence remains uncertain, warranting further research to determine the impact and guide interventions.

## Introduction

In 2022-23 Australia spent $172.3 billion on health care treating disease and injury.^1^ Alcohol and other drug (AOD) use contributes significantly to this burden, accounting for approximately 30,000 hospitalisations and ∼2000 deaths per year.^2^ Residents in very remote areas are ∼11 times more likely to be hospitalised for alcohol-related injuries than those in major cities.^2^ However, the disease burden of major trauma for Australians living in regional to very remote areas is poorly defined.^1, 3^

Several hypotheses have been proposed to explain the influence of AOD use on trauma patients. These include increased risk-taking behaviours, impaired judgement and greater utilisation of healthcare resources.^2, 4^ However, variations in screening across jurisdictions affect the accuracy and comparability of the available data.^4, 5^

Australian and New Zealand health systems define *major trauma* as an Injury Severity Score (ISS) greater than 12.^6^ The ISS is a widely used measure of injury severity that predicts of morbidity, hospital stay and mortality.^7^ Research into AOD-related major trauma has been undertaken across Australia^5, 8–11^ and New Zealand.^12^ Approximately 9% of all major trauma admissions to the Royal Melbourne Hospital involved intoxication, a rate that is increasing and associated with poorer clinical outcomes, including increased mortality in polysubstance users.^9^ Whereas research conducted at the Alfred Hospital found 37.4% of non-transport related major trauma was associated with AOD use.^10^ A prospective study of trauma calls in Christchurch reported that 36.6% of this cohort had evidence of recreational AOD use.^12^ International studies have reported up to 66% of violence related traumatic injuries were linked to AOD use.^13^ In

Queensland (QLD), the QUANTEM report^14^ evaluated alcohol related harm within nightlife precincts. It highlighted that during late night weekend hours, alcohol related traumatic injuries presenting to Emergency Departments (EDs) were significant statewide. There was notable variation was described between metropolitan and regional centres, with highest rates occurring in metropolitan Brisbane. Young males (18-24 years old) were consistently the most overrepresented demographic in alcohol related ED presentations.^14^

To date, no published data exist examining AOD related major trauma in a regional QLD setting. The Townsville University Hospital (TUH) in North QLD is in Outer Regional Australia (Australian Statistical Geography Standard - Remote Area 3; Modified Monash Model category 2).^15^ Townsville University Hospital receives the highest volume of severely injured trauma patients of any hospital outside a metropolitan area in Australia and New Zealand.^16^ It is the sole major trauma centre for all regions north of Mackay and west to the Northern Territory border, and is positioned as a hub to provide definitive trauma care to regional, remote and very remote communities across an ∼800,000km^2^ catchment spanning five Hospital and Health Services in North, Far North and North West QLD.^17^ As the largest tertiary hospital in northern Australia, it also provides specialist referral services for the 700,000 people living throughout this extensive region. The significant challenges associated with the emergency retrieval and care of trauma patients in this expansive area have been well documented.^18, 19^

### Definitions

For this audit, AOD was defined as the presence of any non-clinically administered substance on toxicology screening performed at intake or disclosed in clinical documentation. Polysubstance AOD use was defined as the concurrent detection or use of two or more substances including alcohol. Non-road ambulance transport to TUH was defined as aeromedical, police and private transport. Discharge alternative includes subacute / rehabilitation and correctional facilities. Across EDs in the region, drug and alcohol testing of major trauma patients is not standardised and is generally conducted only when clinically indicated or legally required.

Any injury sustained on a local, state or nationally maintained road network, encompassing all mechanisms of injury (e.g. driver/passenger of motor vehicle, cyclist, pedestrian) was classified as road transport related.

## Material and Methods

Following approval for ethical exemption from the Townsville Hospital and Health Service Audit, Quality and Innovation Review Panel (AQUIRE)^20^ (THHSAQUIRE1681), a retrospective audit was conducted on all major trauma patients with ISS >12 in the Trauma Registry Information System (TRIS) at TUH 1 October 2022-31 October 2023.

Patients were included if they:

- Presented with trauma related injury or died as a result of injury
- Had an ISS >12

Patients were excluded in line with Australian New Zealand Trauma Registry criteria,

- Presentation > 7 days post injury
- Isolated neck of femur fracture (not meeting *major trauma* criteria)
- Injuries directly caused by underlying pathology (for example seizure or stroke)
- >65 years with superficial injuries or co-existing medical conditions that precipitated injury or death (e.g. malignancy, heart failure, renal failure)

Patients were classified into AOD and non-AOD cohorts according to toxicology results, where available, or based on AOD use disclosed in the clinical documentation within the electronic medical record (ieMR). AOD patients were further categorised as demonstrating polypharmacy use. During the data cleaning process, patients were removed from the AOD group if the positive presence of drugs of potential abuse on a toxicology screen was attributed to clinically documented administration by prehospital or ED staff prior to testing.

### Data collection and Analysis

#### Data extraction process

A comprehensive ieMR review was undertaken, including ambulance service records, pathology and toxicology results, triage and nursing notes, ED medical notes, admission notes, Intensive Care Unit (ICU) record and specialist consultation and review notes. Demographic data was sourced from the TRIS.

Descriptive statistics summarised demographics, mechanism of injury, whether transport was involved, drug and/or alcohol profile, clinical severity and hospital variables, including ICU, hospital LOS, and clinical outcomes. Analyses were conducted using Microsoft Excel and IBM SPSS Software version 29.0.0.0 (241).

Normality was assessed using histograms, Q-Q plots, and the Shapiro-Wilk test. Continuous variables demonstrated significant departures from normality (Shapiro-Wilk p < 0.05), with skewness and kurtosis values exceeding acceptable limits (±2). Non-parametric tests were employed for continuous data comparisons, with the Mann-Whitney U test used to compare age, hospital and ICU length of stay and injury severity scores between AOD and non-AOD cohorts.

Categorical variables were analysed using chi-square tests. Chi-square tests of independence assessed whether the distribution of mechanism of injury, transport mode, ICU admission, and patient outcomes differed between AOD and non-AOD groups. Chi-square goodness of fit tests evaluated whether gender distribution deviated from equal distribution, Statistical significance was set at p < 0.05.

Data extracted included:

- Demographics (age, sex, ethnicity)
- Mechanism of injury.

`o` Road (public road) vs non-road injury
`o` Assault vs unintentional/other
- Mode of transport

`o` Road Ambulance
`o` Non-road ambulance transport (including aeromedical retrieval, police/corrections and other)
- ISS
- ICU and acute hospital length of stay (LOS)
- Acute Mortality
- Discharge disposition
- AOD agent use preceding injury
- AOD agent(s) detected.

`o` Patients with multiple agents were classified as polysubstance use within the AOD cohort

## Results

During the study period, 319 patients were admitted to TUH with major trauma. Of these, 110 patients (34.4%) were classified as AOD related cases, while 209 (65.6%) were classified as non-AOD.

### Demographics

#### Age distribution

Most alcohol and drug related major trauma was observed among younger adults (mean age: 40.7 ± 16.7), most commonly in the range of 20-49yo (57.3%). In contrast, non-AOD trauma was more common among older individuals, particularly those >54 years (48.8%). A Mann-Whitney U test confirmed a statistically significant difference in age (p<0.05), with AOD than non-AOD patients (49.0 ± 24.6).

#### Gender distribution

Males were overrepresented across all groups (77.7% of cohort; 77.0% of non-AOD cohort; 79.1% of AOD cohort; 95.0% of polysubstance subgroup; 75.0% of alcohol only and 78.6% of single substance users). Gender distribution did not differ significantly between AOD and non-AOD cohorts (χ²=0.176, p=0.675). Both groups demonstrated significant male predominance compared to the expected 50/50 split (AOD: p< 0.001; non-AOD: p< 0.001).

#### Substance detection

Among the total cohort, alcohol was the most frequently detected substance (28.5%), followed by cannabis (6.9%), amphetamines (4.4%) and benzodiazepines/other (1.9%). Polysubstance use was identified in 6.3% of all cases, most commonly involving combinations of alcohol with amphetamines (35.0%) or cannabis (25.0%).

#### Mechanism and intent

Intentional injuries, particularly assaults, were significantly more prevalent in the AOD group (23.6% vs 4.3%; OR=6.9, 95% CI: 3.1-15.3, p<0.001). Blunt trauma was the dominant mechanism in both cohorts (95.5% AOD vs 95.7% non-AOD: p=1.000). Road transport trauma accounted for 38.2% of the AOD cases, and 33.0% of non-AOD cases, with no significant difference found between cohorts (p=0.357).

#### Transport to hospital

Transport to TUH was primarily non-road ambulance transport, including aeromedical retrieval (53.3% total cohort). No significant difference in transport modes was found between cohorts (p=0.929).

#### Injury severity and hospital outcomes

There were no significant differences in mean ISS between the AOD group (20.4 ± 8.1), the non-AOD group (20.1±7.6), and polysubstance users (20.4±7.8). Intensive care admission rates were higher among the AOD group (46.4% AOD vs. 35.9% non-AOD), though the difference did not reach statistical significance (p=0.069). Intensive care LOS was comparable across groups (AOD: 6.6±5.9 days; polysubstance: 7.4±5.8 days; non-AOD: 7.3 ±7.4days; p=0.075). Hospital LOS was longer among AOD patients (11.9 ± 13.9 vs 10.4 ± 11.4) however, this difference was not statistically significant (p=0.656).

#### Discharge outcomes

Mortality rates were comparable (AOD: 7.3%; non-AOD: 7.7%: OR=0.9, 95% CI: 0.49-2.3, p=0.902), Amongst survivors, discharge destinations were similar, with approximately half being discharged home (51.0% AOD vs 57.5% non-AOD), and half to alternative care settings including subacute/rehabilitation facilities and correctional institutions (49.0% AOD vs 42.5% non-AOD; p=0.283), Among polysubstance users, 25.0% required prolonged rehabilitation and 45.0% were admitted to ICU.

#### Temporal trends

Non-AOD trauma cases increased sharply from June 2023 to September 2023, suggesting potential seasonal variation, while AOD-related trauma remained stable throughout the year.

## Discussion

This study provides the first detailed description of AOD use among major trauma patients in North QLD. Findings demonstrate a clear association between AOD use and injury mechanism, healthcare resource utilisation and patient complexity in a regional Australian setting. Alcohol was the predominant agent, implicating over one third of the major trauma cases, representing a substantial burden of injury. This rate exceeds most national data, but remains lower than international reports, where up to half of all major trauma patients in North America are intoxicated.^21, 22^ These differences likely reflect variation in screening practices, data collection methods and cultural attitudes towards AOD use.

A key finding is the association between AOD use and assault-related injury, with AOD-positive patients having nearly sevenfold higher odds of presenting with an intentional mechanism of injury, highlighting AOD use as a major contributor to interpersonal violence within the major trauma population. AOD-positive patients were also younger, and more frequently male, consistent with established patterns reported in Australian and international trauma literature.^4, 9, 10, 12, 13, 22–24^ These findings identify a high-risk demographic whom targeted prevention strategies may reduce serious injury and associated health expenditure.

Whilst not statistically significant, AOD use in major trauma had a 10% higher ICU admission rate and longer ICU stay, indicating greater demand on critical care resources. Although not statistically significant, AOD-positive patients had lower rates of direct discharge home and higher utilisation of rehabilitation and subacute services, suggesting increased morbidity and prolonged recovery despite similar mortality rates. Previous studies have reported mixed findings regarding mortality in this population,^25, 26^ suggesting that while AOD use may not directly influence survival, it is strongly associated with greater injury complexity and health service utilisation in major trauma patients.^27^

Case identification of AOD related major trauma may be underestimated, due to reliance on clinical documentation and discretionary toxicology testing. The Australasian College for Emergency Medicine has identified that current data collection methods fail to accurately capture the true impact of AOD-related presentations, resulting in systemic underreporting.^28^ Evidence from Australia and New Zealand demonstrates that prospective AOD testing improves detection rates and highlights the epidemiological significance of AOD use within the major trauma population.^4, 12^ International studies further demonstrate substantial variability in AOD screening practices, indicating that clinician driven testing is prone to bias and may underestimate the true burden of AOD use in trauma patients.^29^ Accurate identification is clinically important, as AOD-positive trauma patients experience significantly higher infection and perioperative complication rates, with morbidity increased by up to 30% despite no observed difference in mortality.^26, 30^ Together, these findings reinforce AOD use as an under-recognised contributor to major trauma related morbidity and support the need for standardised testing and prospective research to more accurately define prevalence and outcomes in a regional trauma populations.^4, 31^

### Limitations

The retrospective design and single-centre scope of this study may limit generalisability. Clinician driven testing may introduce selection bias, while patient interviews are subject to recall and reporting bias. Associations should therefore be interpreted with caution.

### Generalisability and implication for policy and practice

The low rate of toxicology screening highlights a critical gap in current trauma care practices. Routine, standardised screening would provide accurate data on AOD prevalence in major trauma, reduce clinician driven bias, and better inform resource allocation in both the acute and rehabilitative phases of care. Consideration of consistent AOD screening at a national level is warranted.

The stable incidence of AOD-related trauma across the year suggests behavioural and psychosocial factors, rather than seasonal or environmental conditions, are key drivers of injury in this cohort. This underscores the importance of targeted prevention strategies, particularly for young males as effective primary prevention may reduce demand for critical health resources. The seasonal trends observed in overall trauma incidence provides insights to inform resource allocation and planning. Predictive research could help healthcare and policy makers in trauma service planning and allocation.

Townsville University Hospital serves one of the largest and most geographically dispersed catchment areas in Australia, encompassing isolated rural and regional communities, as well as mining and agricultural workforces. Despite this heterogeneity, the observed associations mirror national and international findings, reinforcing the broader relevance and applicability of these results.

## Conclusion

In line with international patterns, AOD use is highly prevalent among major trauma patients in North QLD. It is associated with younger age, intentional injury mechanisms like assault, and greater resource needs, including longer hospital stays, increased morbidity and rehabilitation needs, although mortality appears comparable. Reliance on selective toxicology testing likely underestimates the true burden in this regional setting. To better define and understand outcomes, prospective studies with routine screening are needed. Meanwhile, prevention initiatives and integrated discharge planning remain essential to address the complex needs of this vulnerable population.

## Data Availability

All data produced in the present study are available and will be considered on reasonable request.

## Acknowledgements

Acknowledging Associate Professor Mathew Coleman and Dr Vinay Gangathimmaiah for their expertise and advice in support of this research.

We acknowledge the support of the Townsville University Hospital Trauma Research Program, whose resources and expertise were instrumental in the completion of this study. Their commitment to advancing trauma-related research provided essential guidance and contributed significantly to the quality of this work.

We gratefully acknowledge the Townsville Trauma Registry, whose contributions made this audit possible

## Ethical approval statement

Audit approval was provided by the Townsville Hospital and Health Service Audit, Quality and Innovation Review: (THHSAQUIRE1681) 14 February 2024

---

**Table 1.** Demographics and Clinical Characteristics with Statistical Analysis. Two Main Cohorts: AOD (Alcohol and/or Drug use) vs. Non-AOD | **Subgroup:** Polysubstance AOD is nested within the AOD cohort

| Variable | All (n=319, 100%) | AOD Cohort (n=110, 34.4%) |  | Non-AOD Cohort (n=209, 65.6%) | AOD vs Non-AOD p-value | Odds Ratio | Confidence Intervals |
| --- | --- | --- | --- | --- | --- | --- | --- |
|  | M ± SD (n, %) | AOD Overall (n=110) | Polysubstance † (n=20) | M ± SD (n, %) |  |  |  |
| Demographics |  |  |  |  |  |  |  |
| Gender (Male) | 248 (77.7%) | 87 (79.1%) |  | 161 (77.0%) | 0.675 <sup>+</sup> | - | - |
| Mean Age (years) | 46.14 ± 22.53 | 40.67 ± 16.70 | 36.60 ± 12.45 | 49.02 ± 24.61 | <0.05* | - | - |
| Mechanism of Injury |  |  |  |  |  |  |  |
| Road Trauma | 111 (34.8%) | 42 (38.2%) | 11 (55.0%) | 69 (33.0%) | 0.357 <sup>#</sup> | 1.253 | 0.775-2.027 |
| Non-Road Trauma | 208 (65.2%) | 68 (61.8%) | 9 (45.0%) | 140 (66.9%) |  |  |  |
| Injury Intent |  |  |  |  |  |  |  |
| Assault | 34 (10.7%) | 26 (23.6%) | 4 (20.0%) | 9 (4.3%) | <0.001<sup>+</sup> | 6.878 | 3.092-15.303 |
| Unintentional/Other | 285 (89.3%) | 84 (76.4%) | 16 (80.0%) | 200 (95.7%) |  |  |  |
| Trauma Type |  |  |  |  |  |  |  |
| Blunt | 305 (95.6%) | 105 (95.5%) | 19 (95.0%) | 200 (95.7%) | 1.000 <sup>\$</sup> | 0.945 | 0.309-2.892 |
| Penetrating / Burn / Other | 14 (4.4%) | 5 (4.5%) | 1 (5.0%) | 9 (4.3%) |  |  |  |
| Transport Mode |  |  |  |  |  |  |  |
| Road Ambulance | 149 (46.7%) | 51 (46.4%) | 17 (85.0%) | 98 (46.9%) | 0.929 <sup>#</sup> | 0.979 | 0.616-1.555 |
| Non Road Ambulance Transport | 170 (53.3%) | 59 (53.6%) | — | 111 (53.1%) |  |  |  |
| Outcomes |  |  |  |  |  |  |  |
| Mortality | 24 (7.5%) | 8 (7.3%) | — | 16 (7.7%) | 0.902 <sup>#</sup> | 0.946 | 0.392-2.285 |
| Discharge Home | 163 (51.1%) | 52 (51.0%) | 11 (55.0%) | 111 (57.5%) | 0.283 <sup>#</sup> | 0.768 | 0.475-1.244 |
| Discharge alternative | 55 (48.9%) | 50 (49.0%) | 5 (25.0%) | 82 (42.5%) |  |  |  |
| **ICU & Hospital Stay** |  |  |  |  |  |  |  |
| ICU Admission | 126 (39.5%) | 51 (46.4%) | 9 (45.0%) | 75 (35.9%) | 0.069 <sup>#</sup> | 1.544 | 0.966-2.470 |
| Mean ICU LOS (days) | 7.00 ± 6.85 | 6.65 ± 5.94 | 7.36 ± 5.80 | 7.23 ± 7.38 | 0.075 * | - | - |
| Mean Hospital LOS (days) | 10.92 ± 12.32 | 11.95 ± 13.94 | 10.92 ± 12.32 | 10.38 ± 11.36 | 0.656 * | - | - |
| **Injury Severity** |  |  |  |  |  |  |  |
| Mean ISS | 20.40 ± 7.79 | 20.4 ± 8.11 | 20.40 ± 7.79 | 20.11 ± 7.63 | 0.265 * | - | - |

† **Polysubstance AOD:** Cases where more than one substance was detected. This is a subgroup nested within the AOD cohort (n=20 out of n=110 AOD cases).
**LOS:** Length of stay
— **(dash):** Denotes suppressed values where cell counts were <5 or where reporting would permit indirect identification of individuals.
\*: Mann-Whitney.
+: Chi Square Goodness of Fit test.

**Table 2.** Substance Profile and Screening.

| **Outcome** | **Variable** | **AOD cohort**<br>**(n= 110, 34.4% of total cohort)**<br>**(n, % of total cohort)** | **Identified through toxicology screening †**<br>**(n=34, 30.9% of AOD cohort)**<br>**(n, % of AOD cohort)** |
| --- | --- | --- | --- |
| Substance Profile | Alcohol | 91 (28.5%) | 18 (16.3%) |
|  | THC | 22 (6.9%) | 12 (10.9%) |
|  | Amphetamines | 14 (4.4%) | 6 (5.4%) |
|  | Benzodiazepines/Other | 6 (1.9%) | — |
|  | Polysubstance Use ‡ | 20 (6.3%) | 9 (8.1%) |

† The remaining AOD cohort were identified through chart review.
‡ Defined as the presence of more than one substance at hospital presentation.
— (**dash**): Denotes suppressed values where cell counts were <5.

**Figure 1.**
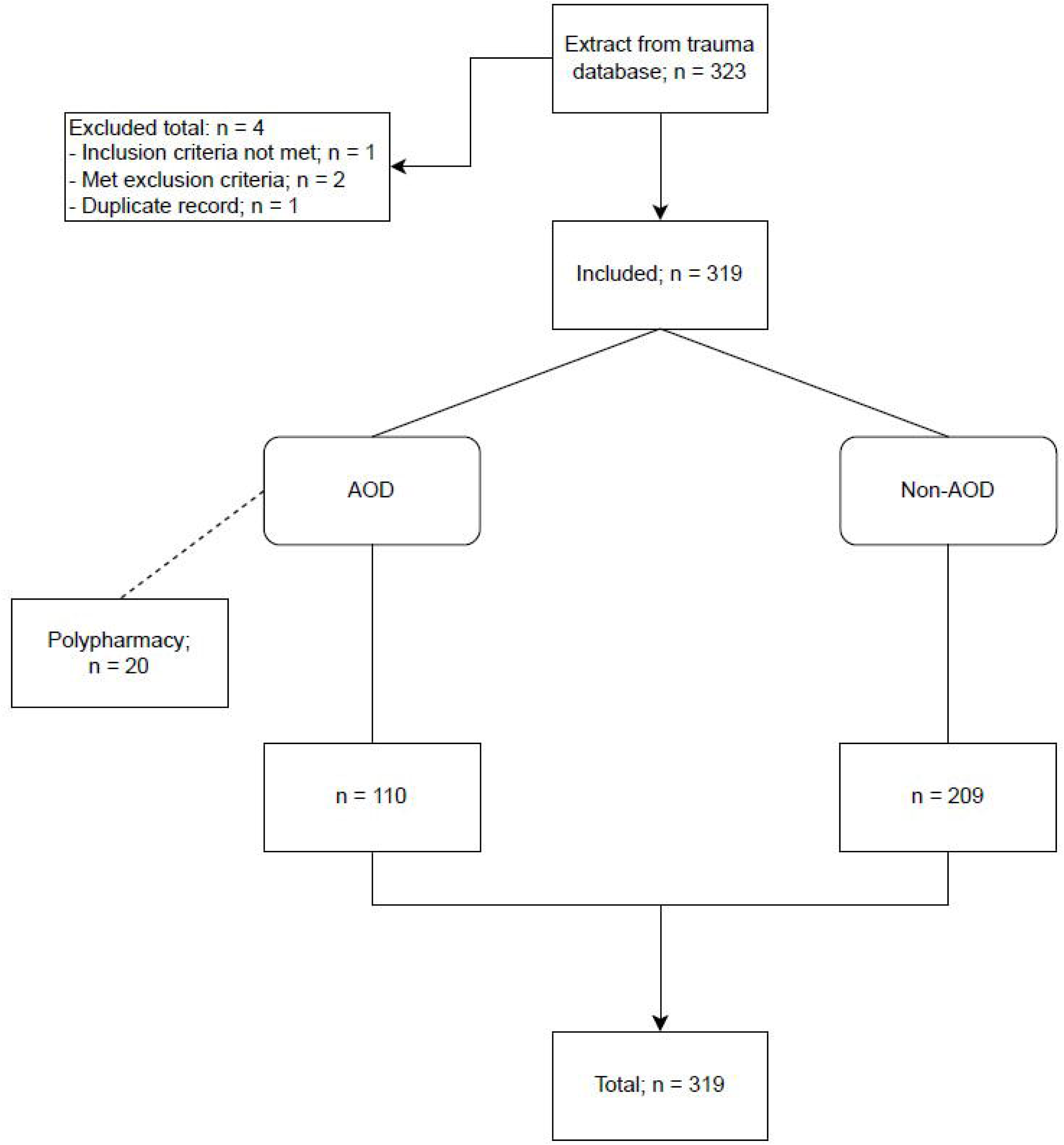
Cohort Allocation Diagram

**Figure 2.**
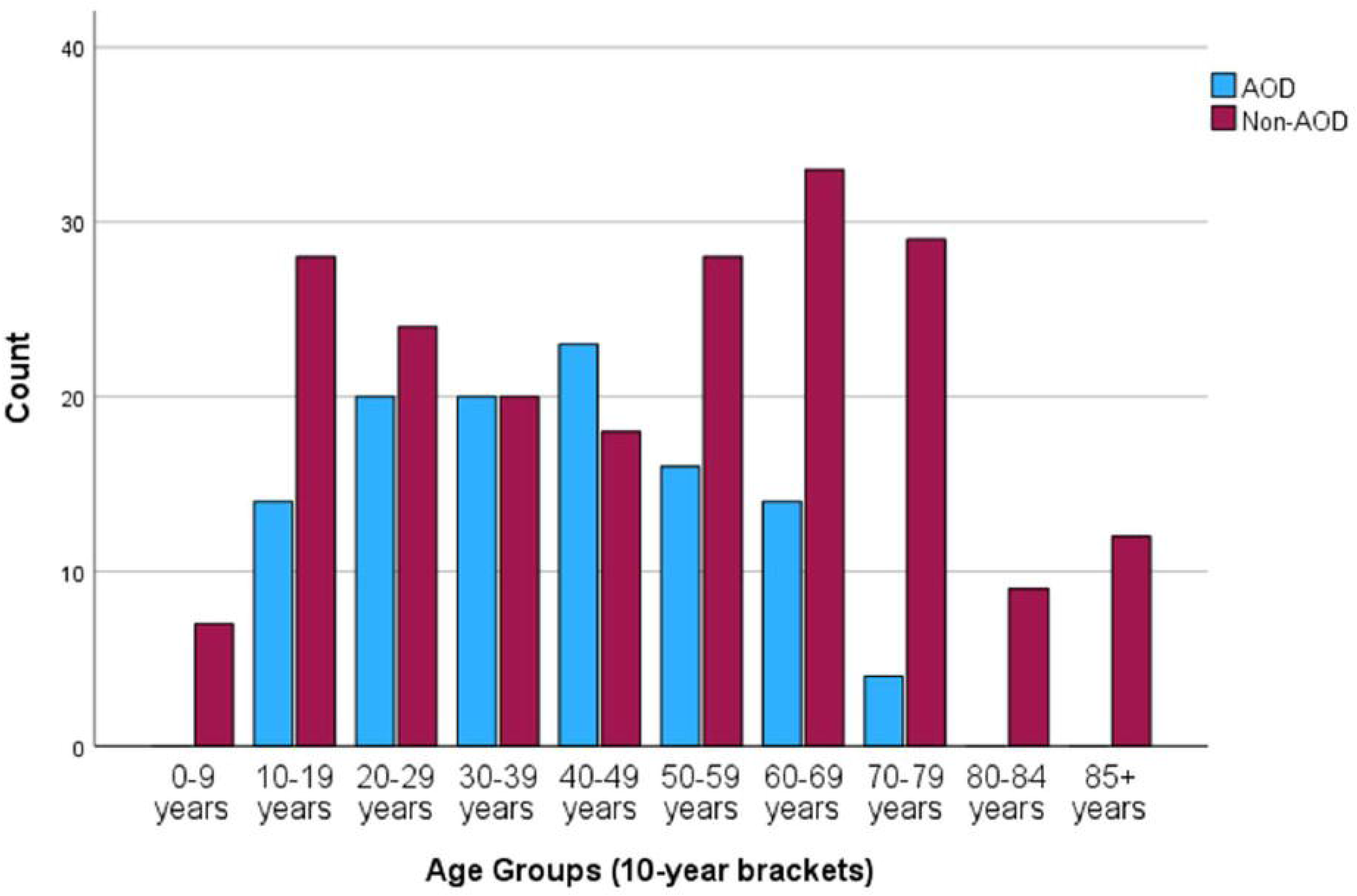
Distribution of age groups by drug and alcohol use

**Figure 3.**
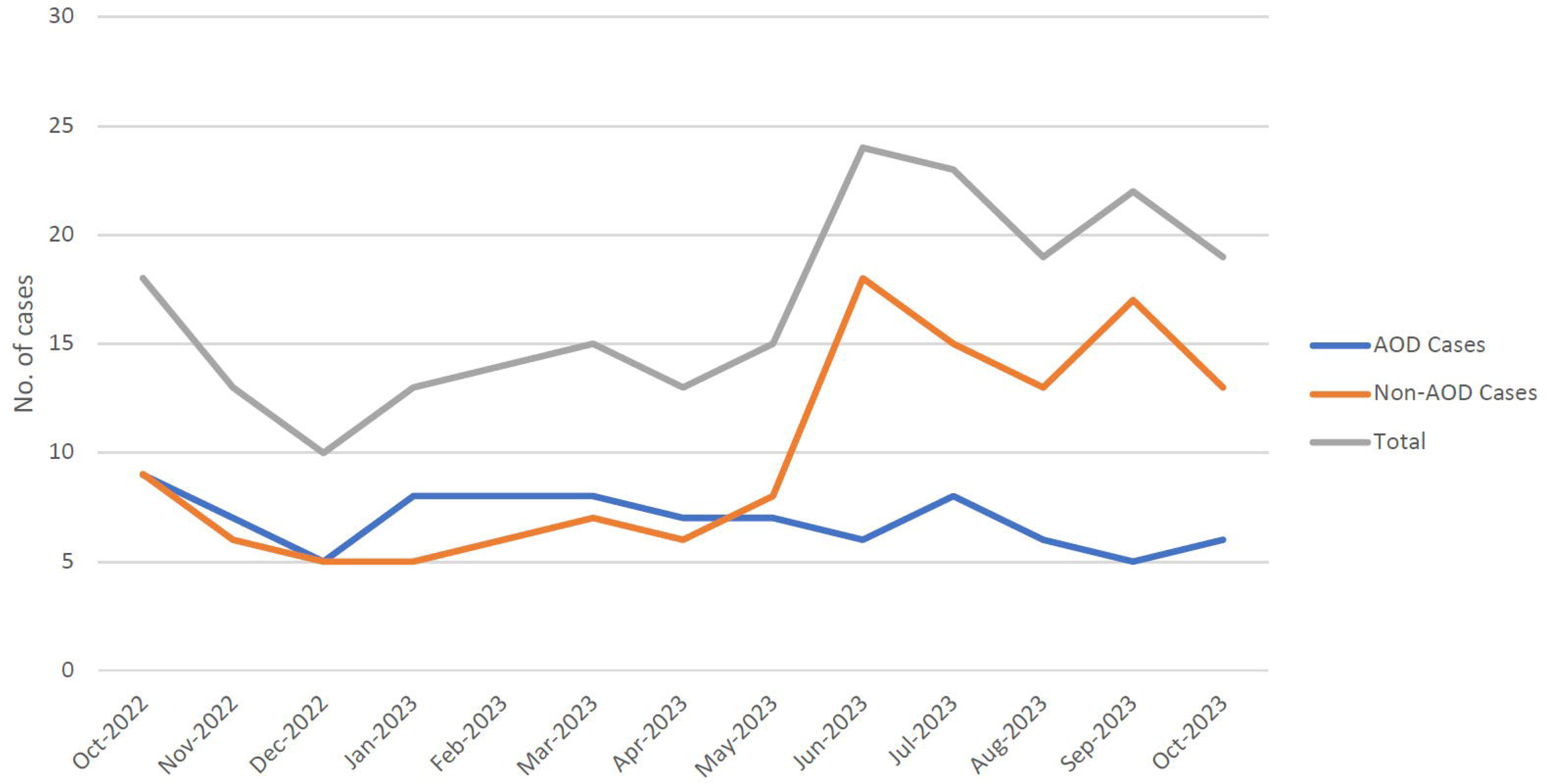
Monthly trauma cases by category from 1 October 2022 to 31 October 2023

